# Accuracy of telephone triage for predicting adverse outcome in suspected COVID-19: An observational cohort study

**DOI:** 10.1101/2021.06.24.21259441

**Authors:** Carl Marincowitz, Tony Stone, Peter Bath, Richard Campbell, Janette Turner, Madina Hussein, Richard Pilbery, Benjamin Thomas, Laura Sutton, Fiona Bell, Katie Biggs, Frank Hopfgartner, Suvodeep Mazumdar, Jennifer Petrie, Steve Goodacre

## Abstract

**Objective:** To assess accuracy of telephone triage in identifying patients who need emergency care amongst those with suspected COVID-19 infection and identify factors which affect triage accuracy.

**Design:** Observational cohort study

**Setting:** Community telephone triage in the Yorkshire and Humber, Bassetlaw, North Lincolnshire and North East Lincolnshire region.

**Participants:** 40, 261 adults who contacted NHS 111 telephone triage services provided by Yorkshire Ambulance Service NHS Trust between the 18^th^ March 2020 and 29^th^ June 2020 with symptoms indicating possible COVID-19 infection were linked to Office for National Statistics death registration data, hospital and general practice electronic health care data collected by NHS Digital.

**Outcome:** Accuracy of triage disposition (self-care/non-urgent clinical assessment versus ambulance dispatch/urgent clinical assessment) was assessed in terms of death or need for organ support at 30, 7 and 3 days from first contact with the telephone triage service.

**Results:** Callers had a 3% (1, 200/40, 261) risk of adverse outcome. Telephone triage recommended self-care or non-urgent assessment for 60% (24, 335/40, 261), with a 1.3% (310/24, 335) risk of subsequent adverse outcome. Telephone triage had 74.2% sensitivity (95% CI: 71.6 to 76.6%) and 61.5% specificity (61% to 62%) for adverse outcomes at 30 days from first contact. Multivariable analysis suggested some co-morbidities (such as chronic respiratory disease) may be over-estimated as predictors of adverse outcome, while the association of diabetes with adverse outcome may be under-estimated. Repeat contact with the service appears to be an important under recognised predictor of adverse outcomes with both 2 contacts (OR 1.77 95% CI: 1.14 to 2.75) and 3 or more contacts (OR 4.02 95% CI: 1.68 to 9.65) associated with clinical deterioration when not provided with an ambulance or urgent clinical assessment.

**Conclusion:** Patients advised to self-care or receive non-urgent clinical assessment had a small but non-negligible risk of serious clinical deterioration. The sensitivity and specificity of telephone triage was comparable to other tools used to triage patient acuity in emergency and urgent care. Repeat contact with telephone services needs recognition as an important predictor of subsequent adverse outcomes.

*What is already known on this topic:*

- Telephone triage has been used to divert patients with suspected COVID-19 to self care or for non-urgent clinical assessments, and thereby help mitigate the risk of health services being overwhelmed by patients who require no speficic treatment.
- Concerns have been raised that telephone triage may not be sufficiently accurate in identifying need for emergency care. However, no previous evaluation of accuracy of telephone triage in patients with suspected COVID-19 infection has been completed.

*What this study adds:*

- Patients advised to self care or receive non-urgent clinical assessment had a small but non-negligible risk of deterioration and significant adverse outcomes.
- Telephone triage has comparable performance to methods used to triage patient acuity in other emergency and urgent care settings.
- Accuracy of triage may be improved by better recognition of multiple contact with services as a predictor of adverse outcomes.

## Background

During the COVID-19 pandemic, there was a risk that hospitals could be overwhelmed by patients who did not need specific treatment. UK government pandemic planning predicted that, in the advent of an influenza or similar pandemic, there could be around 750, 000 excess Emergency Department (ED) attendances in the UK.^1 2^ During the H1N1 outbreak in 2009, ED attendances in the USA increased at some centres by 50%.^3^ Increased ED attendances were largely for patients who did not require hospitalisation.^4^

To reduce this risk, from the 18^th^ February 2020 onwards, NHS England advised patients with suspected infection to contact the NHS 111 service instead of attending health care providers.^5^ NHS 111 is a national, free to use 24 hour telephone triage service for urgent health problems. Initial triage is carried out by trained, non-clinical call advisors using the NHS Pathways clinical decision support software. The endpoint (disposition) is advice on what to do next, in terms of which service to access and the timeframe that this access should occur. If appropriate, the call can be passed onto a clinician (usually a nurse or paramedic) for further assessment and, depending on local arrangements, callers can speak to other specialist clinicians or appointments can be made with relevant services, including general practitioners.

In the first 6-months of the COVID-19 pandemic, ED attendances in the UK decreased by approximately 25%, probably in part due to displacement of care.^6^ Patients who did attend the ED with suspected COVID-19 infection were high acuity with a mortality rate of 15.5%, with lower acuity patients likely being managed via NHS 111.^7^ Indeed, there were almost 3 million NHS 111 calls made across England in March 2020; a record number and double the number of the previous year.^8^ To cope with the increase in call volume, a specific telephone triage pathway for patients with suspected COVID-19 infection was introduced in early February 2020, which underwent rapid updates as the pandemic progressed. Local NHS 111 services used interim triage methods while awaiting implementation of new telephone triage pathways and, due to excess demand, calls started to be diverted to a national centre on the 4^th^ of March.

Telephone triage services provided by NHS 111 appear to have played an important role in the risk assessment of patients with suspected COVID-19 infection in the UK during the pandemic. However, concerns have been raised that telephone triage may miss patients with serious illness, leading to calls for an inquiry into the effectiveness of NHS 111 to identify those in need of medical intervention.^9 10^ There has been no previous evaluation of the accuracy of the clinical risk-assessment performed by this service or, to our knowledge, other telephone triage services for patients with suspected COVID-19 infection.

Our study aimed to:

1. To assess how accurately NHS 111 telephone services identified those who suffered an adverse outcome needing an emergency response.
2. To identify any factors that may have affected the accuracy of telephone triage.

## Methods

### Study Design

The PRIEST study was piloted as the Pandemic Influenza Triage in the Emergency Department (PAINTED) study, part of the UK National Institute for Health Research (NIHR) portfolio of studies to be activated in an influenza pandemic. ^11^ However, it was adapted in February 2020 in response to the COVID-19 pandemic, to include an expanded range of respiratory infections and evaluate pre-hospital urgent and emergency care triage services. This evaluation of NHS 111 telephone services is an observational cohort study that forms part of the PRIEST Study.

### Setting

Yorkshire Ambulance Service NHS Trust (YAS) provides 24-hour emergency and health care services for the Yorkshire and Humber, Bassetlaw, North Lincolnshire and North East Lincolnshire region in the north of England; an area of approximately 6,000 square miles and with a population of 5.3 million. In 2018/19, YAS received more than 998,500 emergency (999) and 1,632,514 NHS 111 calls.

### Data Sources and linkage

Yorkshire Ambulance Service provided a dataset of NHS 111 calls, triaged using an assessment pathway indicating possible COVID-19 infection, received between the 18th March 2020 and 29th June 2020. All patients within the English National Health Service (NHS) are allocated a unique identification number, the NHS number. Records with no NHS number (less than 2%) were not provided as these records could not be associated with a traceable individual without manual review. The dataset consisted of patient identifiers, demographic data, call details and triage dispositions extracted from routinely collected electronic NHS 111 call records.

Patient identifiers were provided to NHS Digital for them to trace the identities of our cohort (i.e. indicate different sets of identifiers belonging to the same patient) and to supply additional individual level demographic, co-morbidity, and outcome data. NHS Digital manages national health and care data collections from a variety of settings and providers in England.^12^ NHS Digital identified records in their collections belonging to patients in our cohort and provided data on patient demographics, limited COVID-related general practice (GP) records, emergency department attendances, hospital inpatient admissions, critical care periods, and death registrations from the Office of National Statistics.

Both YAS and NHS digital removed records belonging to patients who had registered an NHS national data opt-out. The study team excluded patients who had opted out of any part of the PRIEST study and those with inconsistent records (e.g. multiple deaths recorded or death before latest activity). Patient identifiers across all datasets were replaced with a consistent pseudo-identifier to enable the identification of records belonging to individual patients across datasets without revealing patient identifiers.

### Inclusion criteria

Our final cohort consisted of all adult (16+) patients at time of first call (index contact) within the YAS NHS 111 calls dataset who were traced by NHS Digital and for whom a final triage disposition, and therefore urgency of recommended triage, was recorded for their index contact.

### Patient characteristics

Consistent with methods used to estimate the Charlson comorbidity index from these routine data, comorbidities were included if recorded 12 months before the index contact with NHS 111.^13 14^ Immunosuppressant drug use only contributes to the immunosuppression co-morbidity if recorded in the 30 days before index contact. Pregnancy status was based on GP records recorded in the previous 9 months. Frailty in patients older than 65 years was derived from the latest recorded (if any) Clinical Frailty Scale score present in the electronic GP records prior to index contact.^15^ Patients under the age of 65 were given a score of 1, since the score is not validated in this age group. Smoking status was similarly derived from GP records based on the latest recorded (if any) smoking status prior to the index contact.

### Outcome

The primary outcome was death, renal, respiratory, or cardiovascular organ support (identified from death registration and critical care data) at 30 days from index contact.

The secondary outcome was death or organ support at 3 and 7 days from index contact.

### Analysis

We conducted a descriptive analysis of patient demographics, co-morbidities and call disposition and used multivariable logistic regression modelling to confirm known patient characteristics associated with the primary adverse outcome in COVID-19 infection. Obesity was excluded from multivariable analysis due to an observed implausible protective association with the primary outcome which we believe to be an artefact of how these data were collected and recorded in the electronic GP data set. Ethnicity was also excluded from multivariable analysis due to the high proportion of missing data (approximately 25%).

To assess how accurately NHS 111 identified patients with adverse outcomes, the call disposition categories of the index contact were divided into a binary classification of either: ambulance dispatched or other urgent clinical assessment required; and self-care or non-urgent assessment (Supplementary Material 1). Advice and call disposition provided by NHS 111 can change over successive calls as a patient’s condition changes. Therefore, to assess if deterioration was recognised over multiple calls, a sensitivity analysis was conducted in patients who had an adverse outcome in which the disposition of the call immediately before the adverse outcome was used for binary classification.

We assessed the accuracy of the binary triage classification (ambulance dispatch/urgent clinical assessment versus self-care/non-urgent assessment) in terms of sensitivity, specificity, positive predictive value (PPV) and negative predictive value (NPV) for the primary outcome with 95% confidence intervals (CI). To assess whether the implementation of different COVID-19 related NHS Pathways affected the accuracy of triage, accuracy was estimated for the whole study period and in two distinct time periods. The first time period (18/03/2020 to 02/06/2020) encompassed the use of Pathways 19.3.3/4/5/7 by YAS and the second period (02/06/2020 (10:30AM) to 29/06/2020) Pathways 19.3.8/9 which incorporated loss of taste or smell as a feature of COVID-19 infection (Supplementary Material 2).

Patient characteristics of false negatives (those advised to self-care/non-urgent assessment who experienced the primary outcome) and true positives (those provided with an ambulance/urgent assessment who experienced the primary outcome) were compared. In patients with the adverse outcome multi-variable logistic regression was used to identify patient characteristics associated with false negative triage. We also compared the characteristics of false positives (those provided with an ambulance/urgent assessment and not conveyed to hospital) and true negatives (those advised to self-care/non-urgent assessment) among those who did not experience the primary composite adverse outcome. We used multivariable logistic regression to identify factors which predicted false positive triage. Frailty was excluded from multivariable modelling due to a high proportion of missing data (approximately 40% of false negatives).

### Ethical Approval

The North West—Haydock Research Ethics Committee gave a favourable opinion on the PAINTED study on 25 June 2012 (reference 12/NW/0303) and on the updated PRIEST study on 23rd March 2020, including the analysis presented here. The Confidentiality Advisory Group of the NHS Health Research Authority granted approval to collect data without patient consent in line with Section 251 of the National Health Service Act 2006. Access to data collected by NHS Digital was recommended for approval by its Independent Group Advising on the Release of Data (IGARD) on 11^th^ September 2021 having received additional recommendation for approval for access to GP records from the Profession Advisory Group (PAG) on 19^th^ August 2021.

### Patient Public Involvement

The Sheffield Emergency Care Forum (SECF) is a public representative group interested in emergency care research.^16^ Members of SECF advised on the development of the PRIEST study and two members joined the Study Steering Committee. Patients were not involved in the conduct of the study.

## Results

All totals presented are rounded to the nearest 5, with small numbers suppressed to comply with NHS Digital data disclosure guidance.

### Study population

Figure 1 and Table 1 summarise study cohort derivation and the characteristics of the 40, 261 included individuals. In total, 1, 200 people (3%, 95% CI:2.8% to 3.2%) experienced the primary outcome (death or organ support) within 30 days following first contact with telephone triage services and 670 (56%) of adverse outcomes occurred within 7 days of contact. In our study cohort, 8, 165 patients (20.3%, 95% CI:19.9% to 20.7%) were conveyed or self-presented to the ED and 4, 490 (11.2%, 95% CI:10.9% to 11.5%) were admitted as hospital inpatients within 30 days of index contact.

**Table 1:**
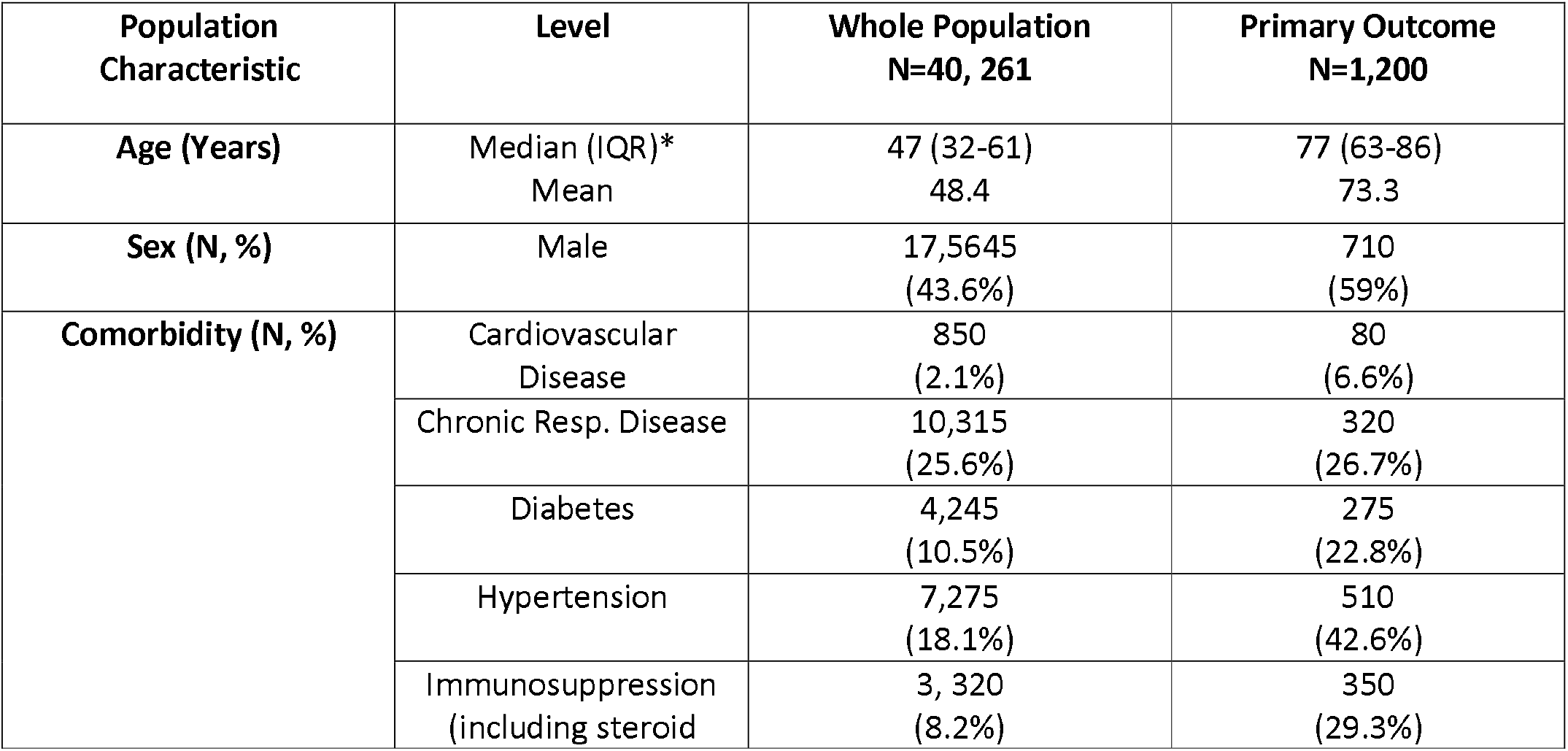
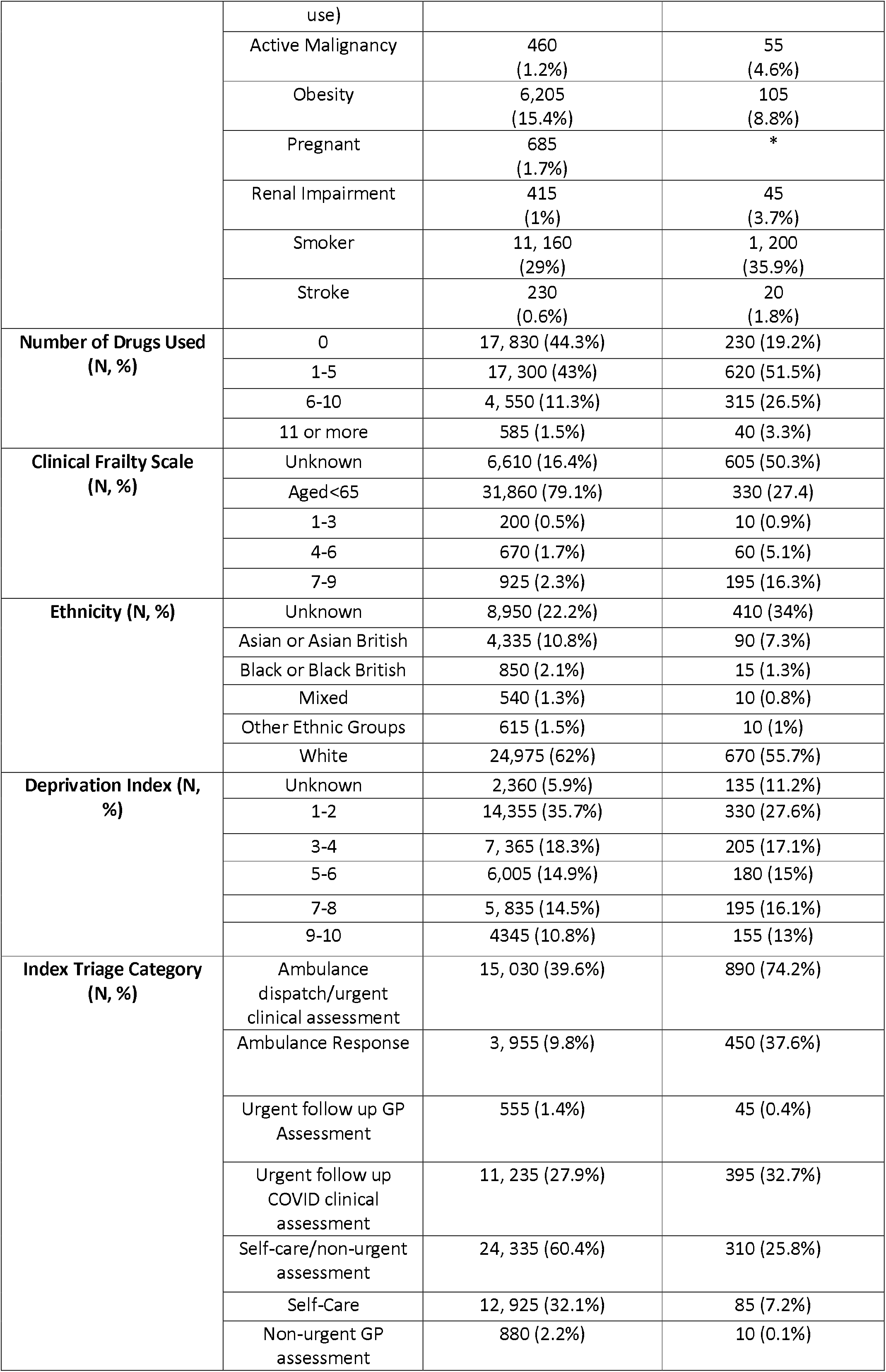
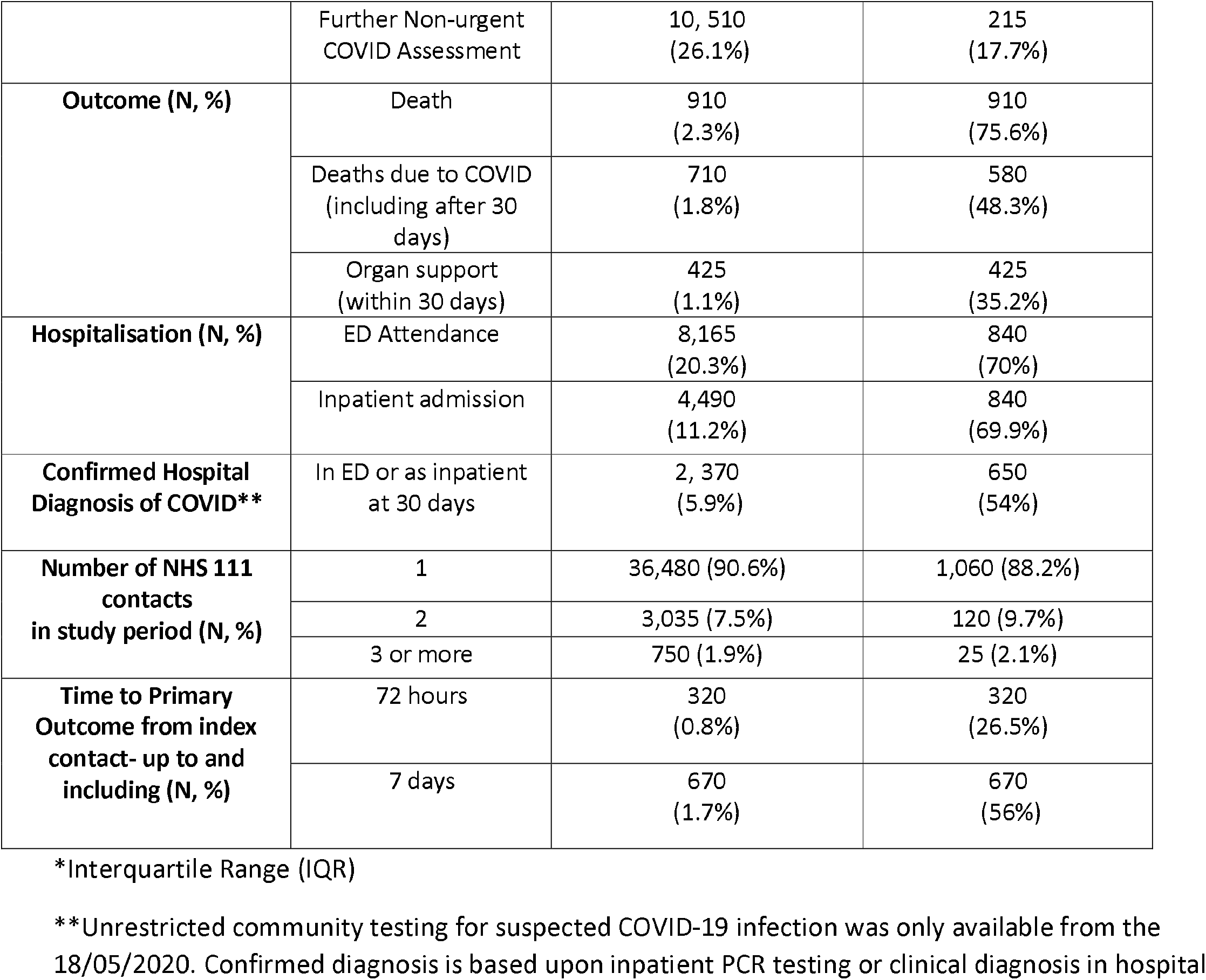
Population characteristics.

**Figure 1:**
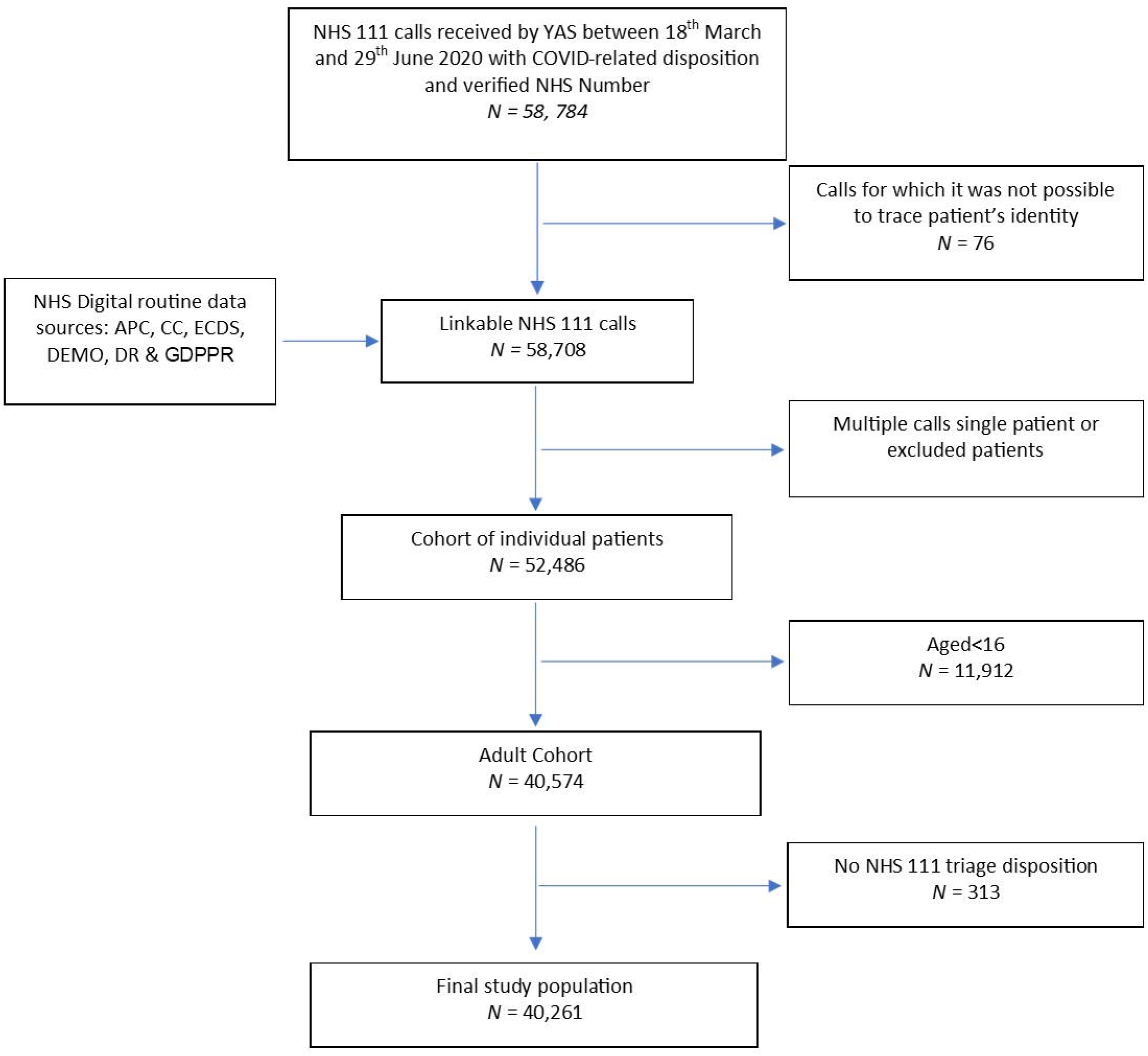
STROBE flow diagram of study population selection

The median age of the whole cohort was 47 (IQR 32-61), the cohort had a higher proportion of females (56.4%) and had high rates of comorbidity (chronic respiratory disease 25.6%, diabetes 10.5% and hypertension 18.1%). In multivariable modelling (Supplementary Material 3), known predictors of adverse outcomes including increasing age, male sex, diabetes and frailty were associated with an increased risk of the primary composite adverse outcome.

### Accuracy of NHS 111 triage

Table 2 shows the accuracy of the binary triage disposition (ambulance dispatch/urgent clinical assessment versus self-care/non-urgent assessment) for the composite primary adverse outcome for the whole study period and two different periods of NHS 111 COVID clinical assessment pathway implementation. Supplementary Material 4 shows the accuracy of the binary triage disposition for the adverse outcomes at seven and three days. A triage disposition of ambulance dispatch/urgent clinical assessment achieved a sensitivity of 74.2% (95% CI: 71.6 to 76.6%) to the primary outcome. If advised to self-care/non-urgent clinical assessment, the chance of experiencing an adverse outcome was approximately 1% (NPV: 98.7%, 95% CI: 98.6 to 98.9%). For patients who contacted NHS 111 multiple times, classification of the triage disposition on the basis of the last call before the primary outcome, instead of index contact, did not noticeably affect these estimates (sensitivity 77.3%, 95% CI: 74.8 to 79.6% and (NPV: 98.9%, 95% CI: 98.7 to 99%).

**Table 2:** Performance of binary NHS 111 triage (ambulance or urgent assessment 4 hours or less) for composite outcome (death or organ support)

| **Adverse outcome up to 30 days (3%, 2.8-3.2%)** |  |  |  |
| --- | --- | --- | --- |
| N=40, 261 | **Adverse Outcome** | **No Adverse Outcome** |  |
| Ambulance/urgent assessment | 890 | 15, 035 | Sensitivity 74.2% (71.6-76.6%)<br>Positive Predictive Value 5.6% (5.2 - 6%) |
| Self-care/ non-urgent assessment | 310 | 24, 025 | Specificity 61.5% (61% - 62%)<br>Negative Predictive Value 98.7% (98.6 - 98.9%) |

| **Time period 1 18/3/20-2/06/2020 (3.1%, 2.9-3.2%)** |  |  |  |
| --- | --- | --- | --- |
| N=36, 124 | **Adverse Outcome** | **No Adverse Outcome** |  |
| Ambulance/urgent assessment | 807 | 13, 079 | Sensitivity 73.2% (70.4-75.7%)<br>Positive Predictive Value 5.8% (5.4 – 6.2%) |
| Self-care/ non-urgent assessment | 296 | 21, 942 | Specificity 62.7% (62.1% - 63.2%)<br>Negative Predictive Value 98.7 (98.5 - 98.8%) |

| **Time period 2 2/06/2020-30/06/2020 (2.4%, 1.9-2.9%)** |  |  |  |
| --- | --- | --- | --- |
| N= 4,137 | **Adverse Outcome** | **No Adverse Outcome** |  |
| Ambulance/urgent assessment | 84 | 1, 957 | Sensitivity 85.7% (76.9-91.7%)<br>Positive Predictive Value 4.1% (3.3 – 5.1%) |
| Self-care/ non-urgent assessment | 14 | 2, 082 | Specificity 51.5% (50% - 53.1%)<br>Negative Predictive Value |
|  |  |  | 99.3 (98.9 - 99.6%) |

Sensitivity of triage disposition was higher for adverse outcomes at three days from index contact (81.4% 95% CI: 76.6 to 85.5%). Specificity was comparable for adverse outcomes at 30 days (61.5% 95% CI: 61 to 62%) and three days (60.8% 95% CI: 60.2 to 61.3%). In the later period of NHS 111 clinical assessment pathway implementation, sensitivity to adverse outcomes at 30 days increased (85.7% 95% CI: 76.9 to 91.7%) but this was associated with a reduction in specificity (51.5% 95% CI: 50 to 53.1%).

### Prediction of false negative or false positive triage

Table 3 compares the characteristics of patients who experienced the primary outcome, and either were (true positives) or were not (false negatives) provided with an ambulance or other urgent clinical assessment. In both groups, approximately 50% of people experienced the primary adverse outcome within seven days of first contact, although a higher proportion of true positives experienced the adverse outcome within three days of contact. Multivariable modelling (Supplementary Material 5), showed that younger age (OR 0.99, 95% CI 0.98 to 1.00), multiple contacts (2 contacts, OR 1.77 95% CI: 1.14 to 2.75 and 3 or more contacts, OR 4.02 95% CI: 1.68 to 9.65), and diabetes (OR 1.66, 95% CI 1.13 to 2.45) were associated with increased risk of false negative triage. The effect estimates for multiple NHS 111 contacts were similar if the triage disposition of last call before the primary outcome (2 contacts, OR 1.96 95% CI: 1.11 to 3.48 and 3 or more contacts, OR 7.78 95% CI: 1.02 to 59.43) was used to classify true positives and false negative.

**Table 3:**
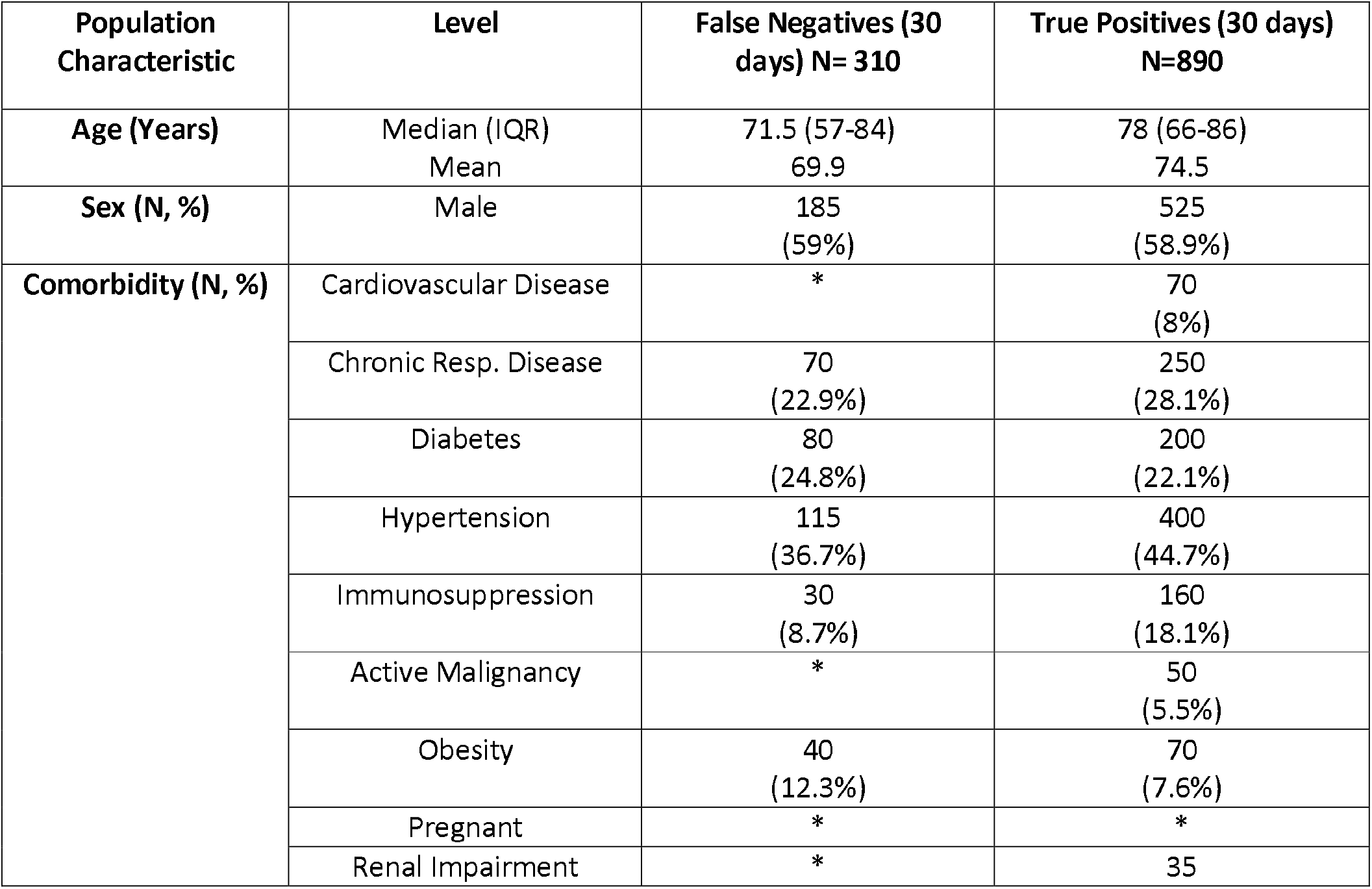
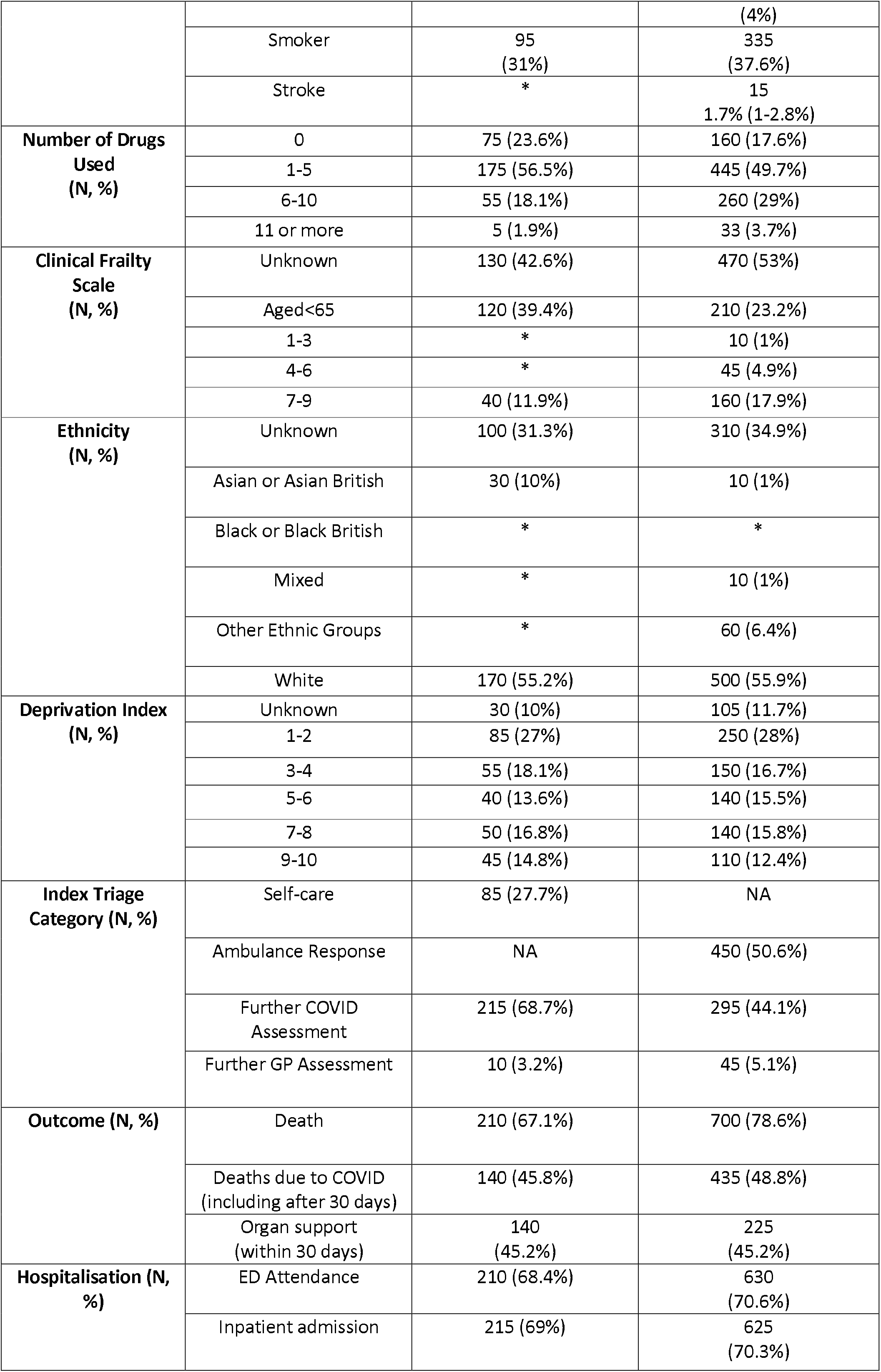
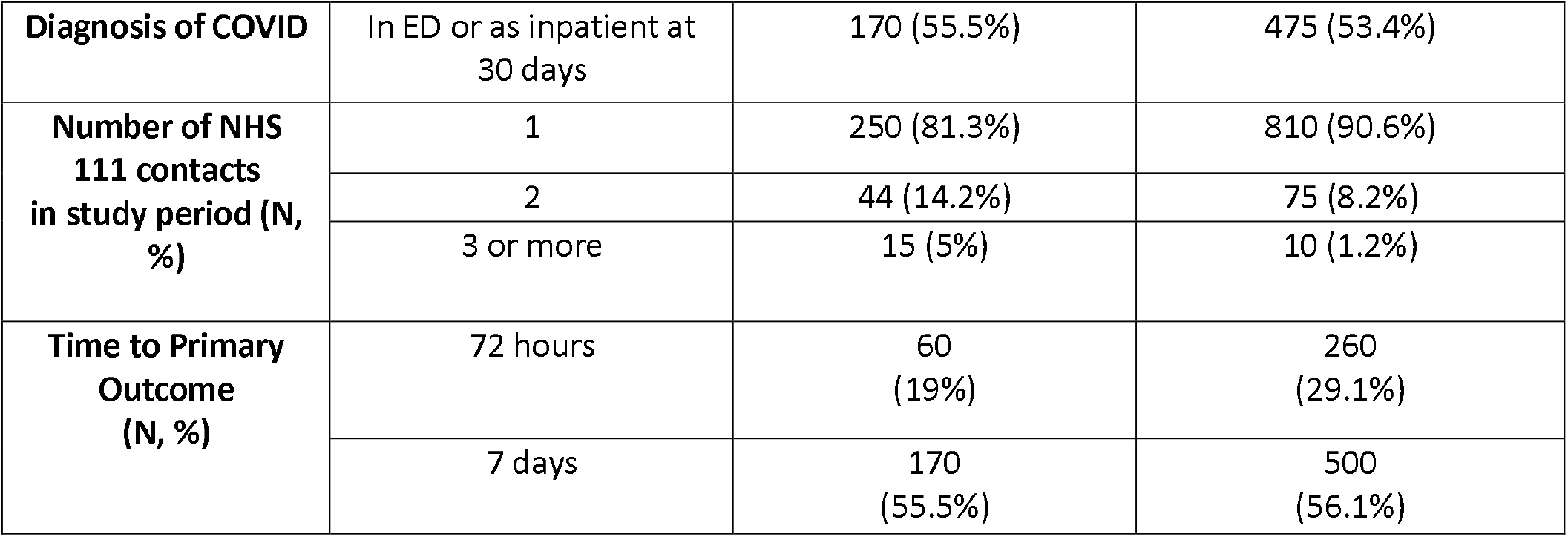
Comparison of False Negatives and True positives.

Table 4 compares the characteristics of patients without the primary outcome who were, either provided with an ambulance/ugent clinical and not conveyed to hospital (False Positives), or not provided with ambulance/urgent clinical response (True Negatives). 24.9% of the cohort were false positives and Supplementary Material 6 presents the results of multivariable modelling to identify factors associated with being a False Positive. Increased risk of being a false positive was most strongly associated with chronic renal impairment (OR1.52 95% CI 1.17 to 1.97), immunosuppression (OR 1.49 95% CI 1.36 to 1.64) and chronic respiratory disease (OR1.31 95% CI 1.22 to 1.40. Weaker predictors included older age, smoking, increased medication use and female sex.

**Table 4:** Comparison of False Positives and True Negatives.

| **Population Characteristic** | **Level** | **False Positives (30 days)**<br>**N= 10, 000** | **True Negative (30 days)**<br>**N=24, 025** |
| --- | --- | --- | --- |
| **Age (Years)** | Median (IQR)<br>Mean | 49 (33-65)<br>49.9 | 44 (31-57)<br>45.2 |
| **Sex (N, %)** | Male | 4,190<br>(41.9%) | 10,460<br>(43.5%) |
| **Comorbidity (N, %)** | Cardiovascular Disease | 235<br>(2.4%) | 310<br>(1.3%) |
|  | Chronic Resp. Disease | 3,160<br>(31.6%) | 5,125<br>(21.3%) |
|  | Diabetes | 1,120<br>(11.2%) | 2,100<br>(8.7%) |
|  | Hypertension | 2,010<br>(20.1%) | 3,320<br>(13.8%) |
|  | Immunosuppression | 1070<br>(10.7%) | 1,278<br>(5.3%) |
|  | Active Malignancy | 120<br>(1.2%) | 150<br>(0.6%) |
|  | Obesity | 1,660<br>(16.6%) | 3,490<br>(14.5%) |
|  | Pregnant | 190<br>(1.9%) | 470<br>(2%) |
|  | Renal Impairment | 135<br>(1.4%) | 140<br>(0.6%) |
|  | Smoker | 3,245<br>(32.4%) | 6,200<br>(25.8%) |

|  | Stroke | 70<br>(0.7%) | 85<br>(0.35%) |
| --- | --- | --- | --- |
| **Number of Drugs Used**<br>**(N, %)** | 0 | 3, 900 (39%) | 12, 230 (50.9%) |
|  | 1-5 | 4, 500 (45%) | 9, 830 (40.9%) |
|  | 6-10 | 1, 420 (14.2%) | 1, 800 (7.5%) |
|  | 11 or more | 190 (1.9%) | 165 (0.7%) |
| **Clinical Frailty Scale**<br>**(N, %)** | Unknown | 1,965 (19.6%) | 2,790 (11.6%) |
|  | Aged<65 | 7,470 (74.7%) | 20,725 (86.3%) |
|  | 1-3 | 60 (0.6%) | 85 (0.4%) |
|  | 4-6 | 225 (2.2%) | 220 (0.9%) |
|  | 7-9 | 290 (2.9%) | 210 (0.9%) |
| **Ethnicity**<br>**(N, %)** | Unknown | 2,100 (21%) | 5,480 (22.8%) |
|  | Asian or Asian British | 1,025 (10.3%) | 2,790 (11.6%) |
|  | Black or Black British | 190 (1.9%) | 555 (2.3%) |
|  | Mixed | 110 (1%) | 355 (1.5%) |
|  | Other Ethnic Groups | 130 (1.3%) | 420 (1.7%) |
|  | White | 6,445 (64%) | 14,420 (60%) |
| **Deprivation Index**<br>**(N, %)** | Unknown | 650 (6.5%) | 1, 180 (4.9%) |
|  | 1-2 | 3, 570 (38.2%) | 8, 730 (38.2%) |
|  | 3-4 | 1, 770 (19%) | 4, 500 (19.7%) |
|  | 5-6 | 1, 470 (15.7%) | 3, 665 (16%) |
|  | 7-8 | 1, 435 (15.4%) | 3, 440 (15.1%) |
|  | 9-10 | 1, 105 (11.8%) | 2, 510 (11%) |
| **Index Triage Category (N, %)** | Self-care | NA | 12, 840 (53.4%) |
|  | Ambulance Response | 1, 645 (16.5%) | NA |
|  | Further COVID Assessment | 7, 935 (79.3%) | 10, 300 (42.9%) |
|  | Further GP Assessment | 360 (3.6%) | 490 (2%) |
| **Number of NHS 111 contacts in study period (N, %)** | 1 | 9,230 (92.3%) | 21,855 (91%) |
|  | 2 | 645 (6.5%) | 1,740 (7.3%) |
|  | 3 or more | 130 (1.3%) | 430 (1.8%) |

## Discussion

### Summary

Our study showed that users of the NHS 111 service who were identified with possible COVID-19 infection had a low (3%) risk of adverse outcome. Telephone triage recommended self-care or non-urgent assessment for the majority (60%), with a very low but non-negligible risk of adverse outcome (1.3%). The sensitivity (74.2%, 95% CI: 71.6 to 76.6%) and specificity (61.5%, 95% 61% to 62%) of telephone triage to the composite primary outcome is similar to that reported for clinical tools used to triage patient acuity in the ED, at a point on the ROC curve with an equivalent balance of sensitivity and specificity.^17^ It is also higher than the reported accuracy of telephone triage for patients with suspected cerebral vascular accidents.^18^ Sensitivity of telephone triage was higher for outcomes at three and seven days (Supplementary Material 4), and sensitivity appeared to be increased at the expense of specificity in the later period of clinical assessment pathway implementation (Table 2).

We used multivariable analysis to identify predictors of false negative and false positive triage. The findings need cautious interpretation, given the limited information available during telephone triage, but suggest that some co-morbidities (such as chronic respiratory disease) may be over-estimated as predictors of adverse outcome, while the association of diabetes with adverse outcome may be under-recognised. Perhaps most striking, is that multiple contacts with NHS 111, in which possible COIVD-19 infection was identifed, was associated with false negative assessment, suggesting that repeat contacts require a more urgent response.

### Strengths and limitations

Although telephone triage has been recommended and widely used during the pandemic in the UK to risk assess patients with suspected COVID-19 to limit potential spread of infection, this appears to be the first evaluation of accuracy.^19^ We have used a large cohort of patients identified from routinely collected telephone triage records and linked this to nationally collected, patient-level healthcare records to provide robust outcome data. We have assessed performance in a cohort of patients with suspected infection which, in the absence of accurate universally available rapid COVID-19 diagnostic tests, reflects the population which urgent and emergency care services must clinically triage. Unrestricted community testing for those with symptoms suggestive of COVID-19 infection was only available from 18/05/2020 and therefore it is not possible to estimate the proportion of confirmed infections. However, known factors associated with adverse outcomes in COVID-19 infection were found to be predictive of the primary outcome in our cohort including increasing age, male sex, diabetes and frailty.^20–22^

Due to the use of routinely collected data there were high rates of missing data for some variables, for example, ethnicity and frailty, which prevented inclusion in some analyses. We have also assumed that if co-morbidities were not recorded in the previous 12-months they are not present. The mechanism of how data are collected and recorded in the routine data sets used means that, as identified for obesity, there may be bias in the classification of patients. The estimated prevalence of obesity in our cohort is 15% (half that reported in the national health survey) and, as weight is not comprehensively and consistently measured by GPs, the observed protective association is likely to reflect unknown characteristics associated with a measurement being taken, rather than obesity itself.^23^

We have evaluated the performance of NHS 111 telephone triage as implemented by the Yorkshire Ambulance Service NHS Trust. Although NHS 111 Pathways software algorithms are developed nationally, there may be variability in local implementation which may affect accuracy. During the study period, calls were diverted between regions and to a national centre due to excess demand. The basis on which calls were selected for diversion is not transparent, but it is possible that patients with less complex health care needs were diverted to the national centre, potentially affecting the generalisability of our results. Our study period includes multiple pathway iterations but due to how rapidly assessment pathways were updated it was not possible to assess the accuracy of individual assessment pathways (Supplementary Material 2). A national online assessment tool was implemented from the end of February 2020 and this may have affected the characteristics of the population using telephone triage services for advice.^24^ However, it was not until June 2020, that the public were advised to use the NHS 111 Online coronavirus service before calling NHS 111.

### Implications

Telephone triage performed comparably to triage methods used for patient acuity in the ED and, given the limited information available, including a lack of physiological parameters, this may reflect the best accuracy that could be achievable.^17 25^ In 2019, the estimated population of Yorkshire and the Humber was 5,502,967 (including children).^26^ On the basis of the number of patients in our cohort and study period, not using telephone triage could have led to around 61 extra ambulances or urgent clinical assessments being provided each day per 1 000, 000 population, without considering diversion to the national centre. Yorkshire Ambulance Service provided a face-to-face response to an estimated 298 incidents per day in March 2020.^27^ Although there was an inevitable degree of misclassification, NHS 111 telephone triage appears to have effectively helped to mitigate the risk of emergency healthcare services being overwhelmed by lower risk patients. This must be weighed against the small but non-negligible risk that patients who were recommended to self-care or have a non-urgent clinical assessment had of serious adverse outcomes. The acceptable risk of deterioration following such triage is subjective and significant variation in risk tolerance between clinicians and public representatives has been demonstrated.^28^ As the pandemic developed, and the risk of emergency services being overwhelmed appeared to reduce, the clinical assessment pathways seemed (arguably rightly) to prioritise sensitivity over specificity. The use of similar telephone triage methods may need to be tailored to the resource constraints and risk tolerance of different health care settings.

Research and information regarding risk factors for adverse outcomes in COVID-19 accumulated rapidly during the initial phase of the pandemic. Early clinical guidelines for the risk stratification of patients with suspected COVID infection, on the basis of previous influenza epidemics, emphasised the importance of respiratory co-morbidities and may have underestimated the risk associated with sex and diabetes.^29^ The results of our multivariable modelling reflects this, with the importance of smoking and chronic respiratory disease appearing to be overestimated and diabetes underestimated. Later NHS clinical guidelines incorporated this evolving research base and emphasised the risk associated with diabetes.^30^ However, the association we found with multiple NHS 111 COVID-19 related contacts and risk of under triage does not appear to have been previously identified and may reflect that patients with repeat contacts represent an unrecognised high risk group. Patients with early representation after discharge from the ED are considered clinically high risk for adverse outcomes and misdiagnosis and this is likely to be reflected in patients who contact NHS 111.^31^

Future research could use our methods for a national evaluation of NHS 111 performance, and to assess regional variation. As there are differences in how providers organise NHS 111 services and how NHS 111 clinical triage pathways were implemented, accuracy may vary. Different models for telephone triage in urgent and emergency care exist internationally.^18 32 33^ NHS 111’s use of trained, non-clinical call advisors for initial assessment contrasts with other national triage services where assessments are performed by nurses and other clinicians.^33^ As as been previously recommended, research is needed to determine the optimal configuration of such services in terms of accuracy and cost-effectiveness.^32^

## Conclusions

We have conducted the first evaluation of accuracy of telephone triage for need for emergency treatment in patients with suspected COVID-19 infection. Telephone triage appears to have had an important role in managing lower-risk patients and potentially preventing many patients who required no specific treatment from attending hospitals or other care providers. This must be weighed against the small but non-negligible risk of serious adverse outcomes in patients advised to self-care or have a non-urgent clinical assessment. Repeat contact with triage services may need more recognition as an important predictor of subsequent deterioration. Future research is needed to determine acceptable risk of deterioration in patients advised to self-care and the optimal configuration of telephone triage services.

## Data Availability

Primary data from this study is not available for sharing from the study team. Data contained from this article is attenable from NHS digital and Yorkshire Ambulance service, following their procedures.

## Authors’ contributions

The idea for the study was conceived by SG, JT, TS, FB, PB and CM. Data processing and linkage was completed by TS and RC. The analyses were completed by CM and MH with specialist statistical advice from SG, JT, LS and PB. All authors contributed to interpretation of results, read and approved the final manuscript.

## Data sharing

The data used for this study are subject to data sharing agreements with NHS digital and Yorkshire Ambulance Service which prohibits further sharing of individual level data. The data sets used are obtainable from these organisations.

