## Supplementary Tables for "Accuracy of telephone triage for predicting adverse outcome in suspected COVID-19: An observational cohort study"

Supplementary Material 1: Classification 111 triage categories

| Ambulance dispatched or other urgent clinical assessment | Self-care or non-urgent assessment |
| --- | --- |
| Ambulance response  Speak to a Clinician from our service Immediately  COVID risk Clinical Assessment service 1 hour  COVID risk Clinical Assessment service 2 hours  COVID risk Clinical Assessment service 4 hours  Speak to a Primary Care Service within 1 Hour  Speak to a Primary Care Service within 2 Hours  Advised to make own way for urgent clinical assessment | COVID Self Care  COVID Coordination Service  COVID risk Clinical Assessment service 6 hours  COVID risk Clinical Assessment service 12 hours  COVID risk Clinical Assessment Service next working day  Home Management  The call is closed with no further action needed  All Dental Dispositions  Any disposition to contact own GP or primary care service  Midwife assessment |

Supplementary Material 2: NHS 111 COVID-19 assessment pathways implementation by Yorkshire Ambulance Service

| NHS 111 Pathway* | Implementation Date | Study Period | Description |
| --- | --- | --- | --- |
| 19.3.3 | 16/3/2020 | 1 | First specific COVID-19 pathway, focus on remote consultations or remote follow-up. |
| 19.3.4 | Not implemented | 1 |  |
| 19.3.5 & 19.3.6 | 03/04/2020 | 1 | Chest pain incorporated as part of COVID assessment pathway  New questions to identify vulnerable Patients |
| 19.3.7 | 10/04/2020 | 1 | New advice provided for signs of deterioration and what to do in patients advised to self-care. |
| 19.3.8 | 02/06/2020 | 2 | More specific triage for symptoms non-specific for COVID e.g. cough or fever offers normal triage plus access to coronavirus triage if certain trigger criteria were met. |
| 19.3.9 | 03/06/2020 | 2 | Incorporates loss of taste or smell as COVID symptom |

*Information regarding specific pathways obtained here <https://digital.nhs.uk/services/nhs-pathways/nhs-pathways-service-information/clinical-release-notes/archived-clinical-release-notes/2020-archived-clinical-release-notes>

Supplementary Material 3: Multi-variable model predicting primary outcome

| **Population Characteristic** | **Level** | **Odds ratio (95% CI)**  **N= 31, 820** |
| --- | --- | --- |
| **Age (Years)** | 1-year increase | 1.06 (1.06 to 1.07) |
| **Sex (N, %)** | Female | 0.48 (0.40 to 0.58) |
| **Comorbidity (N, %)** | Cardiovascular Disease | 0.80 (0.51 to 1.26) |
|  | Chronic Resp. Disease | 0.96 (0.76 to 1.21) |
|  | Diabetes | 1.62 (1.26 to 2.09) |
|  | Hypertension | 1.08 (0.85 to 1.38) |
|  | Immunosuppression  (including steroid use) | 0.97 (0.74 to 1.28) |
|  | Active Malignancy | 1.32 (0.80 to 2.19) |
|  | Obesity | Not included |
|  | Renal Impairment | 1.21 (0.69 to 2.13) |
|  | Smoker | 0.85 (0.69 to 1.04) |
|  | Stroke | 0.54 (0.20 to 1.41) |
| **Number of Drugs Used**  **(N, %)** | 0 | Reference |
|  | 1-5 | 1.03 (0.80 to 1.36) |
|  | 6-10 | 0.93 (0.63 to 1.39) |
|  | 11 or more | 0.87 (0.46 to 1.64) |
| **Clinical Frailty Scale**  **(N, %)** | 1-3 | Reference |
|  | 4-6 | 1.07 (0.71 to 1.61) |
|  | 7-9 | 2.51 (1.74 to 3.61) |
| **Deprivation Index (N, %)** | 1-2 | Reference |
|  | 3-4 | 1.04 (0.80 to 1.34) |
|  | 5-6 | 1.02 (0.77 to 1.34) |
|  | 7-8 | 1.11 (0.85 to 1.45) |
|  | 9-10 | 1.09 (0.81 to 1.46) |
| **Number of 111 contacts**  **in study period (N, %)** | 1 | Reference |
|  | 2 | 1.69 (1.27 to 2.27) |
|  | 3 or more | 2.73 (1.70 to 4.39) |

Supplementary Material 4:

| **Composite Adverse outcome 7 days (1.7%, 1.6-1.8%)** | | | |
| --- | --- | --- | --- |
| N=40, 261 | **Adverse Outcome** | **No Adverse Outcome** |  |
| Ambulance/urgent assessment | 500 | 15,430 | Sensitivity 74.4% (70.9- 77.6%)  Positive Predictive Value  3.1% (2.9 – 3.4%) |
| Self-care/ non-urgent assessment | 170 | 24,160 | Specificity 61% (60.5% - 61.5%)  Negative Predictive Value  99.3% (99.2 - 99.4%) |

| **Composite Adverse outcome 72 hours (0.8%, 0.7-0.9%)** | | | |
| --- | --- | --- | --- |
| N=40, 261 | **Adverse Outcome** | **No Adverse Outcome** |  |
| Ambulance/urgent assessment | 260 | 15,670 | Sensitivity 81.4% (76.6- 85.5%)  Positive Predictive Value  1.6% (1.4 – 1.8%) |
| Self-care/ non-urgent assessment | 60 | 24,275 | Specificity 60.8% (60.3% - 61.3%)  Negative Predictive Value  99.8% (99.7 - 99.9%) |

Supplementary Material 5: Multi-variable model predicting false negatives

| **Population Characteristic** | **Level** | **Odds ratio (95% CI)**  **N= 1, 065** |
| --- | --- | --- |
| **Age (Years)** | 1-year increase | 0.99 (0.98 to 1.00) |
| **Sex** | Female | 1.13 (0.84 to 1.52) |
| **Comorbidity** | Cardiovascular Disease | 0.48 (0.20 to 1.16) |
|  | Chronic Resp. Disease | 1.00 (0.69 to 1.45) |
|  | Diabetes | 1.66 (1.13 to 2.45) |
|  | Hypertension | 0.99 (0.70 to 1.40) |
|  | Immunosuppression  (including steroid use) | 0.62 (0.38 to 1.01) |
|  | Active Malignancy | 0.42 (0.15 to 1.23) |
|  | Obesity | Not included |
|  | Renal Impairment | 0.86 (0.38 to 1.97) |
|  | Smoker | 0.81 (0.58 to 1.12) |
|  | Stroke | 1.99 (0.63 to 6.28) |
| **Number of Drugs Used** | 0 | Reference |
|  | 1-5 | 1.13 (0.74 to 1.74) |
|  | 6-10 | 0.60 (0.33 to 1.10) |
|  | 11 or more | 0.38 (0.11 to 1.27) |
| **Deprivation Index** | 1-2 | Reference |
|  | 3-4 | 1.27 (0.84 to 1.93) |
|  | 5-6 | 1.03 (0.66 to 1.61) |
|  | 7-8 | 1.26 (0.82 to 1.93) |
|  | 9-10 | 1.38 (0.88 to 2.15) |
| **Number of 111 contacts**  **in study period** | 1 | Reference |
|  | 2 | 1.77 (1.14 to 2.75) |
|  | 3 or more | 4.03 (1.68 to 9.65) |

Supplementary Material 6: Multi-variable model predicting false positives

| **Population Characteristic** | **Level** | **Odds ratio (95% CI)**  **N= 32, 195** |
| --- | --- | --- |
| **Age (Years)** | 1-year increase | 1.01 (1.01 to 1.01) |
| **Sex** | Female | 1.05 (1.01 to 1.10 |
| **Comorbidity** | Cardiovascular Disease | 1.04 (0.86 to 1.26) |
|  | Chronic Resp. Disease | 1.31 (1.22 to 1.40) |
|  | Diabetes | 0.84 (0.77 to 0.93) |
|  | Hypertension | 0.97 (0.90 to 1.05) |
|  | Immunosuppression  (including steroid use) | 1.49 (1.36 to 1.64) |
|  | Active Malignancy | 1.25 (0.97 to 1.61) |
|  | Renal Impairment | 1.52 (1.17 to 1.97) |
|  | Smoker | 1.10 (1.04 to 1.16) |
|  | Stroke | 1.23 (0.87 to 1.75) |
| **Number of Drugs Used** | 0 | Reference |
|  | 1-5 | 1.13 (1.06 to 1.21) |
|  | 6-10 | 1.60 (1.42 to 1.81) |
|  | 11 or more | 2.36 (1.82 to 3.07) |
| **Deprivation Index** | 1-2 | Reference |
|  | 3-4 | 0.95 (0.89 to 1.02) |
|  | 5-6 | 0.95 (0.89 to 1.03) |
|  | 7-8 | 0.99 (0.92 to 1.07) |
|  | 9-10 | 1.05 (0.97 to 1.14) |
| **Number of 111 contacts**  **in study period** | 1 | Reference |
|  | 2 | 0.86 (0.78 to 0.95) |
|  | 3 or more | 0.71 (0.58 to 0.88) |
